# Determining important features for dengue diagnosis using feature selection methods

**DOI:** 10.1101/2024.05.05.24306901

**Authors:** Yulianti Paula Bria, Paskalis Andrianus Nani, Yovinia Carmeneja Hoar Siki, Natalia Magdalena Rafu Mamulak, Emiliana Metan Meolbatak, Robertus Dole Guntur

**Affiliations:** Universitas Katolik Widya Mandira, Jl. San Juan No. 1 Penfui Timur, Kabupaten Kupang, Nusa Tenggara Timur 85361, Indonesia; Universitas Nusa Cendana, Jl. Adisucipto Penfui, Kupang, Nusa Tenggara Timur 85001, Indonesia

**Keywords:** Dengue fever, Feature selection, Significant dengue features, Dengue prediction, Dengue diagnosis

## Abstract

**Objectives:** This research aims to determine the important features including symptoms and risk factors for dengue diagnosis.

**Methods:** The dataset for this study is in the form of medical records collected from two hospitals in East Nusa Tenggara Province including Kewapante and Soe hospitals. Feature selection methods including feature importance, recursive feature elimination, correlation matrix from Pearson’s correlation coefficient and KBest were leveraged to determine important features. Important features were also gathered from fifteen Indonesian medical doctors to confirm the results. To obtain the best significant features for dengue prediction, we used six machine learning techniques including logistic regression, k-nearest neighbors, eXtreme gradient boosting, random forests, Naïve Bayes and support vector machines.

**Results:** The random forest classifier yields the highest accuracy for the best combination of features with the accuracy of 0.93 (LR: 0.90 (0.04), KNN: 0.89 (0.04), XGBoost: 0.91 (0.03), RF: 0.93 (0.04), NB: 0.88 (0.09), SVM: 0.89 (0.04)) and precision of 0.90 (LR: 0.86 (0.22), KNN: 0.67 (0.14), XGBoost: 0.77 (0.13), RF: 0.90 (0.13), NB: 0.66 (0.20), SVM: 0.66 (0.18)). This study shows the significant features for dengue diagnosis including fever, fever duration, headache, muscle and joint pain, nausea, vomiting, abdominal pain, shivering, malaise, loss of appetite, shortness of breath, rash, bleeding nose, bitter mouth, temperature and age.

**Conclusions:** This beneficial information can help society in differentiating dengue from non-dengue diseases including malaria, typhoid fever, COVID-19 and other dengue-like symptoms diseases. This is pivotal to educate society to seek medical advice when dengue symptoms appear.

## INTRODUCTION

Dengue infection is a life-threatening disease spread by female mosquitos, *Aedes aegypti*. This disease is one of the most prevalent diseases in many countries including Indonesia. In 2022, Indonesia contributed to 143,266 dengue cases with the mortality rate in the same year with 1,237 people [1]. Based on the report from Ministry of Health of Indonesia in the week-19 2023, Indonesia had 31,380 dengue cases which claimed 246 people [1]. This indicates that dengue eradication must be prioritized by the government and society without ignoring other priority health problems such as tuberculosis, malaria, stunting, etc.

Early-stage dengue diagnosis is challenging since dengue shares similar symptoms to other diseases including malaria, typhoid fever, and even COVID-19. Malaria, for example, shares the same symptoms with dengue fever such as fever, nausea, vomiting and headache [2].

Some countries have their own identified symptoms for dengue fever. Australia, for example, defines the combination of fever, headache, arthralgia, myalgia, rash, nausea and vomiting as dengue symptoms [3]. Whereas, Singapore uses the combination of fever, headache, backache, myalgia, rash, abdominal discomfort and thrombocytopenia for dengue symptoms [3]. In Indonesia, medical doctors refer to Dengue guideline for diagnosis, treatment, prevention and control [4] dan comprehensive guidelines for prevention and control of Dengue and Dengue Haemorrhagic Fever [5]. The guideline for dengue diagnosis and treatment is issued by Ministry of Health of Indonesia, which is used as a reference for medical personnel [6]. This is adopted from WHO dengue case classification [4].

The number of deaths in East Nusa Tenggara (NTT) province from dengue cases in 2022 was 29 out of 3,376 cases [7]. These cases spread all over NTT’s districts. Most of the death cases were because of the severe conditions. People often visit the nearest medical centre when they identify rash or severe conditions because of the lack of knowledge of dengue symptoms and risk factors [8]. Understanding important features of dengue is beneficial to avoid the progression to severe condition, which can avoid death. This information is helpful to seek medical advice as soon as dengue symptoms appear. The important features are pivotal to develop early-stage dengue detection tools to assist in dengue diagnosis from other dengue-like symptoms diseases such as malaria, typhoid fever and even COVID-19.

Even though there are some guidelines used to diagnose and treat dengue [4,5], different countries have different symptoms [3]. Therefore, it is essential to identify first significant symptoms that contribute most for dengue prediction in Indonesia, which will be done in this study. This study will also use the combination of symptoms and dengue risk factors that contribute most for dengue diagnosis.

To obtain significant features from datasets, we use feature selection methods. Feature selection methods are often used to minimize the number of input variables that are considered to be the most significant to a machine learning model to improve the model performance [9,10]. In recent years, numerous publications focus on the implementation of feature selection methods for disease prediction [9–13]. In the classification stage, most researchers use machine learning techniques such as BayesNet [9,10,13], support vector machine [9,11] and tree-based classifiers [9,10,13].

The use of feature selection for dengue fever has been implemented successfully by Ramasami et al. [14]. They focus on applying feature selection process and relative analysis to enhance the performance of dengue prediction models. In Indonesia, dengue prediction research has been focused on the use of machine learning techniques for predicting the dengue outbreak [15], predicting number of dengue incidents [16–18], forecasting model for dengue fever [19], and focusing spatial modelling for dengue fever [20–22]. To the best of our knowledge, this study is the first study to elaborate some feature selection methods to determine significant features for dengue diagnosis based on medical records collected. The results will be compared with the knowledge gathered from the fifteen Indonesian medical doctors’ knowledge to confirm the results. This research also aims to provide important symptoms and factors for malaria diagnosis in Indonesia.

## METHODS

### Flow chart of Study

Figure 1 shows the approach to obtain significant features for dengue diagnosis.

**Figure 1.**
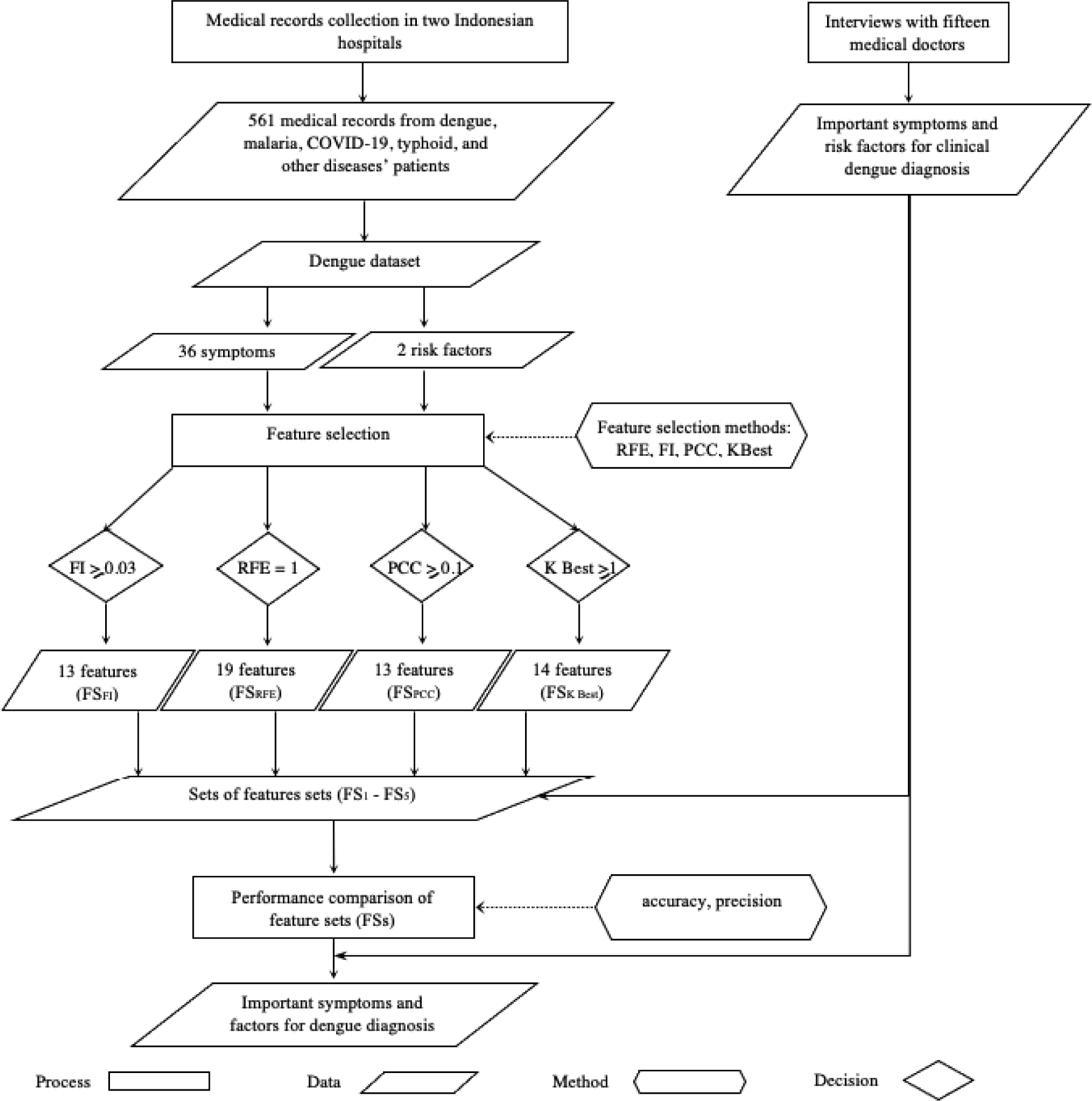
The approach for determining important features for dengue diagnosis.

### Data collection – medical records collection

To obtain the dengue dataset, we conducted the data collection in two hospitals in Kewapante Hospital, Maumere in Sikka District and Soe Hospital in South Central Timor District of NTT Province. Medical records were collected in the department of medical records of each hospital after obtaining the data collection approvals from the hospital directors in each hospital. Medical records of patients diagnosed with dengue fever or other dengue-like symptoms diseases, such as malaria, typhoid fever, COVID-19, dyspepsia, pneumonia, and gastritis, were collected for the years 2017-2023. These two hospitals’ medical records were paper-based, requiring manual recording using an Excel spreadsheet. The features recorded from the medical records collected are shown in the following list:

1. Age;
2. Gender;
3. Temperature;
4. All recorded symptoms;
5. Duration of fever;
6. Working diagnosis;
7. Laboratory test results;
8. Final diagnosis.

Table 1 shows the characteristics of collected medical records from the two Indonesian hospitals. The total medical records collected (*n*) is 561 records. The medical records consist of 473 non-dengue cases and 88 dengue cases. Features in the form of symptoms are indicated using S and features in the form of risk factors are indicated using F. The collected dataset will then be named as a dengue dataset, which has 36 symptoms and two risk factors. Most of the symptoms are binary in the form of 1 for Yes or Female and 0 for No or Male. The duration of fever (S_2_), temperature (S_25_) and age (F_1_) are in the form of number. The target in the dataset is Diagnosis, which is the form of the binary value (1 for dengue and 0 for non-dengue diseases).

**Table 1.** Characteristics of collected medical records (*n*=561).

| Notation | Feature | $n$ | % |
| --- | --- | --- | --- |
| **Symptoms** |  |  |  |
| $S_1$ | Fever | | |
|  | Yes | 318 | 56.68 |
|  | No | 243 | 43.32 |
| $S_2$ | Duration of fever | Mean 2.38, SD 3.62 | |
| $S_3$ | Headache | 158 | 28.16 |
|  | Yes | 403 | 71.84 |
|  | No |  |  |
| S4 | Arthralgia / Myalgia |  |  |
|  | Yes | 24 | 4.28 |
|  | No | 537 | 95.72 |
| S5 | Nausea |  |  |
|  | Yes | 290 | 51.69 |
|  | No | 271 | 48.31 |
| S6 | Vomiting |  |  |
|  | Yes | 196 | 34.94 |
|  | No | 365 | 65.06 |
| S7 | Abdominal pain |  |  |
|  | Yes | 129 | 22.99 |
|  | No | 432 | 77.01 |
| S8 | Shivering |  |  |
|  | Yes | 33 | 5.88 |
|  | No | 528 | 94.12 |
| S9 | Body pain |  |  |
|  | Yes | 23 | 4.10 |
|  | No | 538 | 95.90 |
| S10 | Heartburn |  |  |
|  | Yes | 224 | 39.93 |
|  | No | 337 | 60.07 |
| S11 | Chest pain |  |  |
|  | Yes | 44 | 7.84 |
|  | No | 517 | 92.16 |
| S12 | Dizziness |  |  |
|  | Yes | 171 | 30.48 |
|  | No | 390 | 69.52 |
| S13 | Malaise |  |  |
|  | Yes | 489 | 87.17 |
|  | No | 72 | 12.83 |
| S14 | Loss of appetite |  |  |
|  | Yes | 289 | 51.52 |
|  | No | 272 | 48.48 |
| S15 | Sneezing |  |  |
|  | Yes | 182 | 32.44 |
|  | No | 379 | 67.56 |
| S16 | Coughing |  |  |
|  | Yes | 304 | 54.19 |
|  | No | 257 | 45.81 |
| S17 | Shortness of breath / fast breathing |  |  |
|  | Yes | 140 | 24.96 |
|  | No | 421 | 75.04 |
| S18 | Rash |  |  |
|  | Yes | 10 | 1.78 |
|  | No | 551 | 98.22 |
| S19 | Bleeding nose |  |  |
|  | Yes | 3 | 0.53 |
|  | No | 558 | 99.47 |
| S20 | Bitter mouth |  |  |
|  | Yes | 22 | 3.92 |
|  | No | 539 | 96.08 |
| S21 | Sore throat |  |  |
|  | Yes | 11 | 1.96 |
|  | No | 550 | 98.04 |
| S22 | Blurry vision |  |  |
|  | Yes | 2 | 0.36 |
|  | No | 559 | 99.64 |
| S23 | Seizure |  |  |
|  | Yes | 7 | 1.25 |
|  | No | 554 | 98.75 |
| S24 | Diarrhea |  |  |
|  | Yes | 74 | 13.19 |
|  | No | 487 | 86.81 |
| S25 | Temperature | Mean 37.06, SD 1.93 |  |
| S26 | Sweating |  |  |
|  | Yes | 28 | 4.99 |
|  | No | 533 | 95.01 |
| S27 | Swallowing pain |  |  |
|  | Yes | 27 | 4.81 |
|  | No | 534 | 95.19 |
| S28 | Pale |  |  |
|  | Yes | 25 | 4.46 |
|  | No | 536 | 95.54 |
| S29 | Jaundice |  |  |
|  | Yes | 1 | 0.18 |
|  | No | 560 | 99.82 |
| S30 | Anaemia |  |  |
|  | Yes | 2 | 0.36 |
|  | No | 559 | 99.64 |
| S31 | Black water |  |  |
|  | Yes | 2 | 0.36 |
|  | No | 559 | 99.64 |
| S32 | Constipation |  |  |
|  | Yes | 37 | 6.60 |
|  | No | 524 | 93.40 |
| S33 | Flatulence |  |  |
|  | Yes | 37 | 6.60 |
|  | No | 524 | 93.40 |
| S34 | Feeling anxious |  |  |
|  | Yes | 3 | 0.53 |
|  | No | 558 | 99.47 |
| S35 | Bleeding coughing |  |  |
|  | Yes | 25 | 4.46 |
|  | No | 536 | 95.54 |
| S36 | Loss of consciousness |  |  |
|  | Yes | 5 | 0.89 |
|  | No | 556 | 99.11 |
| **Non-symptom-related factors** |  |  |  |
| F1 | Age | Mean 31.66, SD 24.57 |  |
| F2 | Gender |  |  |
|  | Female | 304 | 54.19 |
|  | Male | 257 | 45.81 |

SD: standard deviation

### Interview results with fifteen Indonesian medical doctors

To confirm the results from the significant features obtained from the feature selection process, we interviewed 15 Indonesian medical doctors about important symptoms and risk factors for dengue diagnosis. These 15 medical doctors were provided with the structured interview questions regarding symptom and risk factors for dengue diagnosis. The questions were in Bahasa Indonesia. Therefore, we translated it in English. Table 2 shows the summarized interview results from the 15 Indonesian medical doctors about symptoms and risk factors for clinical diagnosis of dengue fever.

**Table 2.**
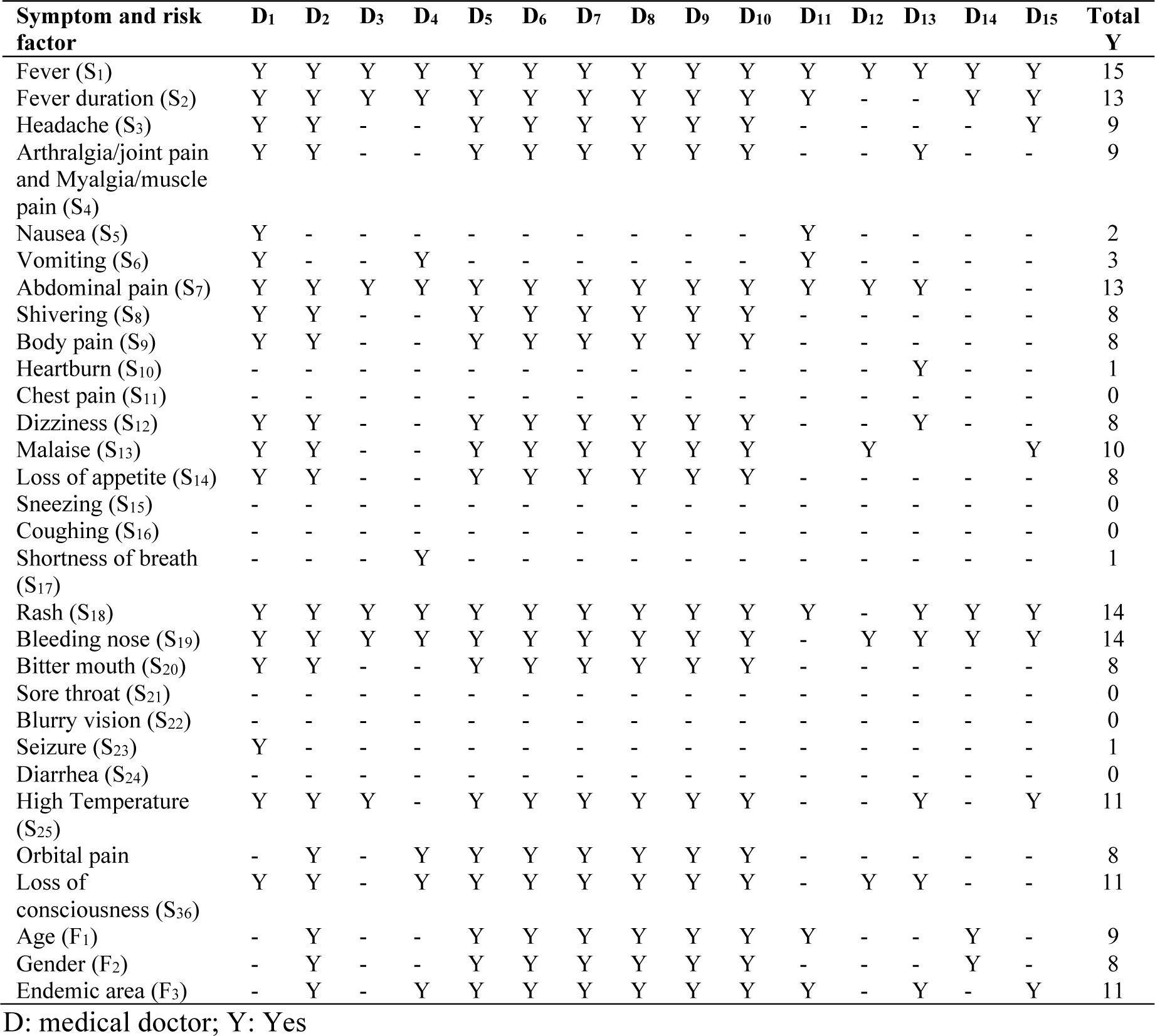
The summarized symptoms and risk factors from the fifteen Indonesian medical doctors.

| Symptom and risk factor | D<sub>1</sub> | D<sub>2</sub> | D<sub>3</sub> | D<sub>4</sub> | D<sub>5</sub> | D<sub>6</sub> | D<sub>7</sub> | D<sub>8</sub> | D<sub>9</sub> | D<sub>10</sub> | D<sub>11</sub> | D<sub>12</sub> | D<sub>13</sub> | D<sub>14</sub> | D<sub>15</sub> | Total Y |
| --- | --- | --- | --- | --- | --- | --- | --- | --- | --- | --- | --- | --- | --- | --- | --- | --- |
| Fever (S<sub>1</sub>) | Y | Y | Y | Y | Y | Y | Y | Y | Y | Y | Y | Y | Y | Y | Y | 15 |
| Fever duration (S<sub>2</sub>) | Y | Y | Y | Y | Y | Y | Y | Y | Y | Y | Y | - | - | Y | Y | 13 |
| Headache (S<sub>3</sub>) | Y | Y | - | - | Y | Y | Y | Y | Y | Y | - | - | - | - | Y | 9 |
| Arthralgia/joint pain and Myalgia/muscle pain (S<sub>4</sub>) | Y | Y | - | - | Y | Y | Y | Y | Y | Y | - | - | Y | - | - | 9 |
| Nausea (S<sub>5</sub>) | Y | - | - | - | - | - | - | - | - | - | Y | - | - | - | - | 2 |
| Vomiting (S<sub>6</sub>) | Y | - | - | Y | - | - | - | - | - | - | Y | - | - | - | - | 3 |
| Abdominal pain (S<sub>7</sub>) | Y | Y | Y | Y | Y | Y | Y | Y | Y | Y | Y | Y | Y | - | - | 13 |
| Shivering (S<sub>8</sub>) | Y | Y | - | - | Y | Y | Y | Y | Y | Y | - | - | - | - | - | 8 |
| Body pain (S<sub>9</sub>) | Y | Y | - | - | Y | Y | Y | Y | Y | Y | - | - | - | - | - | 8 |
| Heartburn (S<sub>10</sub>) | - | - | - | - | - | - | - | - | - | - | - | - | Y | - | - | 1 |
| Chest pain (S<sub>11</sub>) | - | - | - | - | - | - | - | - | - | - | - | - | - | - | - | 0 |
| Dizziness (S<sub>12</sub>) | Y | Y | - | - | Y | Y | Y | Y | Y | Y | - | - | Y | - | - | 8 |
| Malaise (S<sub>13</sub>) | Y | Y | - | - | Y | Y | Y | Y | Y | Y | - | Y | - | - | Y | 10 |
| Loss of appetite (S<sub>14</sub>) | Y | Y | - | - | Y | Y | Y | Y | Y | Y | - | - | - | - | - | 8 |
| Sneezing (S<sub>15</sub>) | - | - | - | - | - | - | - | - | - | - | - | - | - | - | - | 0 |
| Coughing (S<sub>16</sub>) | - | - | - | - | - | - | - | - | - | - | - | - | - | - | - | 0 |
| Shortness of breath (S<sub>17</sub>) | - | - | - | Y | - | - | - | - | - | - | - | - | - | - | - | 1 |
| Rash (S<sub>18</sub>) | Y | Y | Y | Y | Y | Y | Y | Y | Y | Y | Y | - | Y | Y | Y | 14 |
| Bleeding nose (S<sub>19</sub>) | Y | Y | Y | Y | Y | Y | Y | Y | Y | Y | - | Y | Y | Y | Y | 14 |
| Bitter mouth (S<sub>20</sub>) | Y | Y | - | - | Y | Y | Y | Y | Y | Y | - | - | - | - | - | 8 |
| Sore throat (S<sub>21</sub>) | - | - | - | - | - | - | - | - | - | - | - | - | - | - | - | 0 |
| Blurry vision (S<sub>22</sub>) | - | - | - | - | - | - | - | - | - | - | - | - | - | - | - | 0 |
| Seizure (S<sub>23</sub>) | Y | - | - | - | - | - | - | - | - | - | - | - | - | - | - | 1 |
| Diarrhea (S<sub>24</sub>) | - | - | - | - | - | - | - | - | - | - | - | - | - | - | - | 0 |
| High Temperature (S<sub>25</sub>) | Y | Y | Y | - | Y | Y | Y | Y | Y | Y | - | - | Y | - | Y | 11 |
| Orbital pain | - | Y | - | Y | Y | Y | Y | Y | Y | Y | - | - | - | - | - | 8 |
| Loss of consciousness (S<sub>36</sub>) | Y | Y | - | Y | Y | Y | Y | Y | Y | Y | - | Y | Y | - | - | 11 |
| Age (F<sub>1</sub>) | - | Y | - | - | Y | Y | Y | Y | Y | Y | Y | - | - | Y | - | 9 |
| Gender (F<sub>2</sub>) | - | Y | - | - | Y | Y | Y | Y | Y | Y | - | - | - | Y | - | 8 |
| Endemic area (F<sub>3</sub>) | - | Y | - | Y | Y | Y | Y | Y | Y | Y | Y | - | Y | - | Y | 11 |

D: medical doctor; Y: Yes

### Machine learning techniques used

In this study, we employ commonly used machine learning techniques in dengue and malaria prediction including, support vector machine (SVM) [23–25], random forest (RF) [25,24], eXtreme gradient boosting (XGBoost) [23], logistic regression (LR) [23,24], k-nearest neighbour (KNN) [26] to develop dengue classifiers that can accurately distinguishing dengue from non-dengue diseases.

### Performance metrics used

To evaluate the performance of the classifiers, we use two performance metrics including accuracy and precision. The formula for the two performance metrics can be seen in Equations (1)–(2).

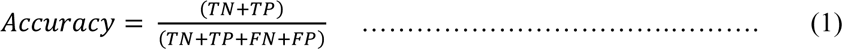

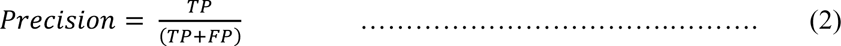

Where,

TP is the number of dengue records that are correctly classified;

TN is the number of non-dengue records that are correctly classified;

FP is the number of non-dengue records classified as dengue;

FN is the number of dengue records classified as non-dengue.

### Feature selection methods used

Feature selection filters redundant or irrelevant features [27]. By reducing the number of features, it will minimize the computational cost of the prediction and increase the performance of the machine learning classifier. The feature selection methods assess the relationship between each feature and the target feature and choose the input features that have the strongest correlation with the target feature [28]. The higher the score, the more the feature is related to the target feature.

In this study, feature selection methods used to determine important features are feature importance [29], recursive feature elimination (RFE) [30,31], correlation matrix from Pearson’s correlation coefficient (PCC) [2,28] and KBest [27].

### Ethical Statement

This research was approved by Human Ethics Committee of Widya Mandira Catholic University (reference number: 001/WM.H9/LPPM/SKKEP/X/2023).

## RESULTS

Table 3 shows the four feature selection scores from FI, RFE, CM and KBest for features that meet the threshold. The threshold value for each feature selection method is used to obtain the most important features from FI (>=0.030), RFE (1), PCC (>=0.100) and KBest (>1.000). From this first process of filtering, some features are eliminated.

**Table 3.**
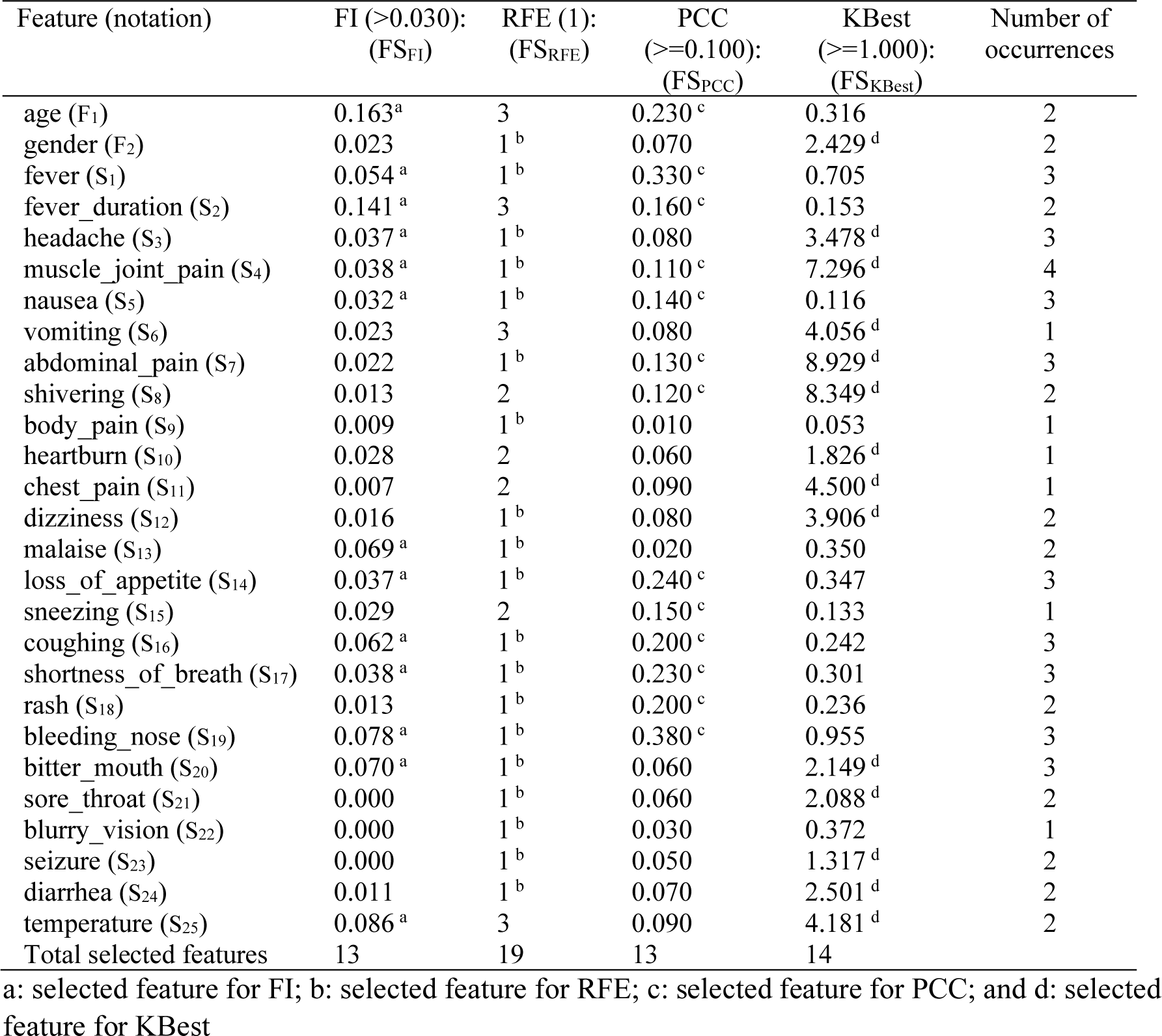
The number of occurrences of features in the four feature selection methods with their selection results.

Table 3 also shows the total number of significant features for each feature selection method. Feature importance from RF has 13 significant features. RFE selects 19 significant features. There are 13 significant features for PCC and 14 significant features for KBest respectively. These results show that each feature selection method has its own combination of significant features. In Colum 6 of Table 3, we total number of occurrences for each feature based on the given thresholds from the four feature selection methods. The higher the number of occurrences, the more significant the feature. There are some features that are significant for three or four feature selection methods. Muscle_joint_pain (S_4_), for example, is the only feature choosed by the four feature selection methods. This indicates that this feature is the most significant feature among other features. From Table 3, we generate FS_1_ from selected features >=3, FS_2_ from selected features >=2 and FS_3_ from selected features >=1. FS_4_ consists of FS_1_ and selected features = 1. FS_5_ consists of FS_PCC_ and the selected symptoms from 15 medical doctors.

In order to choose the significant features for dengue diagnosis based on various combination of features, we compare feature sets (FS_s_) generated. Table 4 shows the performance comparison from various features sets generated. As shown in Table 4, the most stable performance for almost all machine learning classifiers is FS_5_. Therefore, the most significant features for dengue prediction are the combination of features of FS_5_. The random forest classifier yields the highest accuracy for FS_5_ with the accuracy of 0.93 and precision of 0.90.

**Table 4.**
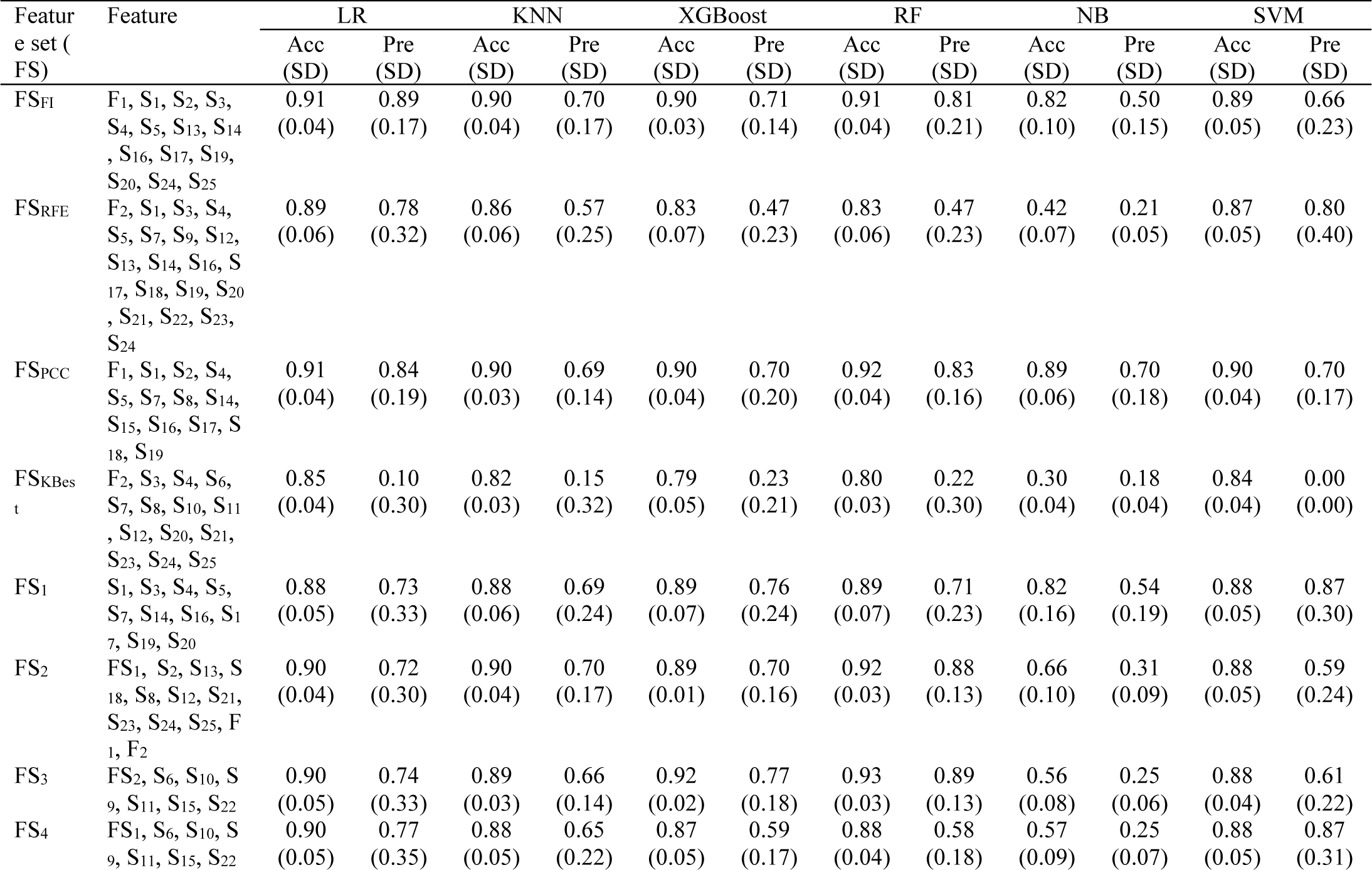
Performance comparison of features sets generated with the standard deviation values.

## DISCUSSION

Table 4 shows that significant features for dengue prediction are fever (S_1_), fever duration (S_2_), headache (S_3_), muscle joint pain (S_4_), nausea (S_5_), vomiting (S_6_), abdominal pain (S_7_), shivering (S_8_), malaise (S_13_), loss of appetite (S_14_), sneezing (S_15_), coughing (S_16_), shortness of breath (S_17_), rash (S_18_), bleeding nose (S_19_), bitter mouth (S_20_), temperature (S_25_) and age (F_1_). However, not all these features are dengue symptoms. It is important to note that the dataset consists of dengue records and other medical records including malaria, COVID-19, dyspepsia, gastritis, typhoid fever and pneumonia. We will discuss which symptoms and risk factors that are important for dengue predictions or dengue diagnosis with the confirmation of medical doctors knowledge.

Fever, fever duration and high temperature are three important dengue symptoms. For fever, three out of four feature selection methods select this symptom as an important feature. All fifteen medical doctors interviewed also agree that one of the most important dengue features is fever. Even though only two feature selection methods including FI and PCC chose fever duration as an important feature, fever normally starts 4-10 days after infection and last for 2-7 days [32]. Based on the medical doctors interviewed and medical records collected, temperature also has a significant contribution for dengue prediction that can reach 39-40°C. Eleven medical doctors interviewed agree that high temperature of fever is important to distinguish dengue from other diseases such as malaria and typhoid fever. Therefore, it is important to include fever, fever duration and high temperature of fever as three important features for dengue diagnosis.

Arthralgia/joint pain and myalgia/muscle pain are two symptoms that are considered as the most significant features for the dengue prediction and dengue diagnosis [33]. All the four feature selection methods indicate these two symptoms are important for distinguishing dengue from other diseases including malaria, typhoid fever, COVID-19, dyspepsia and pneumonia. Nine medical doctors also consider these two symptoms as significant symptoms for dengue diagnosis.

Headache is one of the most important symptoms in diagnosing and predicting dengue [33]). The three feature selection methods other than PCC consider this symptom essential for dengue diagnosis. In addition, it is also confirmed by nine medical doctors interviewed.

Nausea is considered as one of the most significant symptoms for dengue diagnosis [33]. That also applies for vomiting [34]. However, if persistent vomiting occurs then the individual might progress to the severe state [33]. Two medical doctors agree that nausea is part of dengue symptoms whereas three medical doctors agree that vomiting is an important symptom for dengue diagnosis. In the prediction perspective, nausea is more considered significant because it is selected by three feature selection methods. Whereas vomiting is least significant as only KBest selects this symptom. However, these two symptoms are highly correlated, thus it is important to consider both symptoms as dengue symptoms. Loss of appetite is considered a symptom that can indicate individuals suffer from dengue. Eight medical doctors interviewed confirm that this symptom is also considered as a dengue symptom. This symptom is also selected by three feature selection methods other than KBest.

Even though shivering is associated with malaria [2,33], shivering is also important for dengue diagnosis and prediction. Eight medical doctors interviewed also agree that shivering is also a dengue symptom. In the dengue prediction perspective, shivering is also an important feature for dengue prediction as it is selected by two feature selection methods including PCC and KBest as part of significant features.

Malaise is an important symptom for dengue diagnosis and it normally happens when individuals are in severe condition [33]. In addition, ten medical doctors also confirm that this symptom is essential in dengue diagnosis. It is also selected by two feature selection methods including FI and RFE.

Bleeding nose is one of the most important symptoms in dengue diagnosis as part of bleeding manifestations [33,34]. This symptom with other bleeding manifestations indicate that individuals progress is in severe condition. Fourteen medical doctors interviewed agree that to determine an individual suffers from dengue is to check the presence of the bleeding nose. Moreover, three feature selection methods selected this symptom as a significant feature for dengue prediction.

Similar to the bleeding nose, the presence of rashes in skin is also pivotal in distinguishing dengue from other similar diseases such as malaria and typhoid fever [33]. Fourteen medical doctors confirm that a rash in an individual’s body is a distinguishing symptom that led their initial diagnosis to dengue. This symptom is also selected in two feature selection methods including RFE and PCC.

Abdominal pain is considered as one of the dengue symptoms especially when someone in the severe state [33,34]. Thirteen medical doctors also confirm that this symptom is essential to determine dengue from other diseases. This symptom is also selected by three feature selection methods other than FI as a significant symptom for dengue prediction.

Shortness of breath or fast breathing is one of dengue symptoms that indicates the severe state of dengue [33]. This is also confirmed by one medical doctor interviewed. This symptom is also selected by three feature selection methods other than KBest as the important feature for dengue prediction.

Age can be considered as one of the important risk factors for dengue diagnosis [35]. Even though six medical doctors do not consider this factor as an important feature for dengue diagnosis, nine medical doctors include this factor as feature that should not be overlooked when diagnosing potential dengue patients. Two feature selection methods including FI and PCC also consider this factor important for dengue prediction. Normally, individuals younger than 15 years old are prone to dengue infection [36]. Bitter mouth is associated with malaria as this symptom is considered as one of malaria symptoms [37,38]. However, interestingly eight medical doctors interviewed agree that this symptom also can be found in individuals who suffer from dengue. This symptom also appears in three feature selection methods other than PCC. Thus, this symptom should not be ignored when diagnosing potential dengue patients.

From the dengue prediction perspective, sneezing and coughing are important features. Three feature selection methods select this symptom as significant features for dengue prediction. However, no medical doctors confirm that sneezing and coughing are part of dengue symptoms. Sneezing and coughing might be the distinguished symptom to determine COVID-19 from dengue. It is important to know that the dataset consists of medical records from COVID-19 patients. Besides, sneezing and coughing are known as COVID-19 symptom [39,40]. Therefore, sneezing and coughing are important for dengue prediction but not necessarily are dengue symptoms.

This study does not include other features such as orbital pain, history of previous suffering from dengue and history of visiting endemic dengue areas. In this study, all this information were not found in the medical records collected. This opens the room for the future studies. The significant features as results from this study can be used to develop reliable and powerful machine learning techniques, which later can be used to develop early-stage dengue prediction tools.

In conclusion, there are four findings of this study. First, there are 17 symptom features including fever, fever duration, headache, muscle and joint pain, nausea, vomiting, abdominal pain, shivering, malaise, loss of appetite, sneezing, coughing, shortness of breath, rash, bleeding nose, bitter mouth, temperature and one risk factor feature including age that are important for dengue prediction. However, sneezing and coughing are not necessarily important for dengue diagnosis. Second, arthralgia/joint pain and myalgia/muscle pain are the most significant features for the dengue prediction. Third, even though a bitter mouth symptom is highly related to malaria diagnosis, this study suggests that the medical doctors should not ignore the bitter mouth symptom in diagnosing dengue as this symptom is also important for dengue prediction. Fourth, random forest classifier yields the most stable performance for dengue prediction. Knowledge of these features are essential to educate society about significant symptoms and risk factors for dengue to avoid progression to severe conditions, which can lead to death. The findings of this study can also be used as a reference for medical doctors in differentiating dengue from non-dengue diseases including malaria, COVID-19 and typhoid fever.

## Data Availability

All data produced in the present study are not available publicly as they are generated from the medical records of patients. However, the unrevealed identity dataset can be provided upon reasonable request to the authors.

## ACKNOWLEDGMENTS

We would like to thank Widya Mandira Catholic University for providing the research grant for this research project. We would also like to express our gratitude to the Director of Kewapante Hospital Sikka, and the Director of Soe Hospital South Central Timor, and to the Department of Permission Affair in Sikka, South Central Timor, and in East Nusa Tenggara Province — Indonesia. We are grateful to the medical records staff and the fifteen medical doctors for their cooperation.

## AUTHOR CONTRIBUTIONS

Conceptualization: Bria YP, Nani PA, Siki YCH, Meolbatak EM, Guntur RD, Mamulak NMR. Data curation: Bria YP, Nani PA, Mamulak NMR. Formal analysis: Bria YP, Siki YCH, Meolbatak EM, Guntur RD. Funding acquisition: Bria YP. Methodology: Bria YP, Nani PA, Siki YCH, Meolbatak EM, Guntur RD, Mamulak NMR. Project administration: Mamulak NMR. Writing – original draft: Bria YP, Nani PA, Siki YCH, Meolbatak EM, Guntur RD, Mamulak NMR. Writing – review & editing: Bria YP, Nani PA, Siki YCH, Meolbatak EM, Guntur RD, Mamulak NMR.

